# Exploring the Digital Landscape: A Comprehensive Analysis of Infertility Narratives on Instagram

**DOI:** 10.1101/2024.04.07.24305110

**Authors:** Abhijay Gillellamudi, Venkata Sujatha Vellanki, Binay Kumar Panjiyar

**Affiliations:** Abhijay Gillellamudi Sree ramachandra medical college porur India

**Author notes:** Corresponding Author: Binay Kumar Panjiyar MBBS GCSRT, PGME, Harvard Medical School.

**Keywords:** Instagram, infertility, health posts, social media

## Abstract

In the digital age, the prevalence of infertility and the trend of delaying childbearing have significantly influenced fertility choices. Social media, particularly Instagram, has emerged as a vital part of daily life for many, with a notable percentage of American adults engaging daily. Recognizing the impact of online platforms, this cross-sectional observational study, conducted in May 2023, delved into the nature and quality of 673 Instagram posts related to infertility. It aimed to analyze the information and support available to individuals’ navigating infertility, particularly focusing on the posts DISCERN and Global Quality Scores (GQS) to assess their reliability and quality.

The findings reveal that many women derive substantial benefits from their online experiences. Instagram serves as a source of coping mechanisms for the challenges associated with infertility diagnosis and treatment. The platform allows for the sharing of personal feelings and experiences, providing a valued sense of community and support. However, it also presents a risk of spreading misinformation, underscoring the need for careful consideration of content quality.

The study emphasizes the importance of accessible and reliable online resources. It suggests that healthcare professionals should play a more active role in guiding patients to trustworthy information. In conclusion, while social media offers valuable support and unique insights for individuals dealing with infertility, the quality and reliability of the information remain critical concerns. The study advocates for a more informed and conscientious use of these platforms, highlighting the potential for both support and misinformation.

## Introduction and Background

The global incidence of infertility, defined as the inability to conceive after 12 months of regular, unprotected intercourse, has been a growing concern, affecting approximately 8-12% of couples worldwide, amounting to 48.5 million couples (1). The increasing trend to delay childbearing, influenced by sociocultural and economic factors, has led to a rise in the average age of first birth, now around 30 years for women in many parts of the world (2). This delay is associated with decreased fecundity and increased risk of complications, thus propelling the conversation around infertility to the forefront of reproductive health discussions (3).

The advent of the digital era, characterized by the rapid diffusion of the internet and, particularly, the proliferation of high-speed (broadband) internet, has revolutionized the way individuals seek and share health-related information (4). Social media platforms, especially Instagram, have become integral to daily routines, shaping public discourse and personal experiences in profound ways. A significant portion of the American population, 67% of young adults aged 18–29 and 47% of adults aged 30-49, are reported to use Instagram daily (5). The platform allows users to post pictures and videos, add captions, and use hashtags to categorize posts, creating a dynamic and interactive information-sharing environment.

The role of social media in health communication is particularly pertinent in the context of infertility. Patients often turn to the internet as their first source of fertility-related information, seeking not just medical facts but also emotional support and personal narratives (6). While the internet offers unparalleled access to information, the quality and reliability of this information can vary dramatically. The potential for misinformation is a significant concern, as inaccurate health information can lead to poor health outcomes, increased anxiety, and misinformed decision-making (7).

The importance of reliable online resources is underscored by the finding that patients often seek the advice of a physician on social media, highlighting the platform’s role in patient-physician interactions (8). Furthermore, health professionals have begun using social media as a tool for public health messaging and medical education, aiming to provide evidence-based information to a broad audience (9). Despite these positive developments, the lack of quality control on social media platforms remains a challenge.

The current study is premised on the understanding that while social media has the potential to provide support and valuable information to individuals facing infertility, the quality and reliability of this information are not guaranteed. By analyzing Instagram posts related to infertility, this study aims to shed light on the nature of the information and support available to individuals navigating this challenging condition. Specifically, the study evaluates the posts based on the DISCERN and Global Quality Scores (GQS), established tools for assessing the quality and reliability of health information (10, 11).

In summary, as the trend of delayed childbearing continues and the use of social media for health-related information grows, understanding the nature and quality of online discourse around infertility is crucial. This study provides a comprehensive analysis of Instagram posts related to infertility, offering insights into the potential benefits and risks associated with seeking health information on social media platforms. The findings aim to inform strategies for improving the quality and reliability of online health information, ultimately enhancing patient care and public health outcomes.

## Materials and Methods

### Study Design and Setting

This cross-sectional observational study was conducted in May 2023 to analyze the nature and quality of information regarding infertility shared on Instagram. This platform was chosen due to its widespread use and significant impact on health-related communication. The study’s protocol was designed to adhere to the principles outlined in the Declaration of Helsinki.

### Data Collection

A systematic search for Instagram posts was conducted using the platform’s search functionality. The search was limited to posts made public and in the English language. Posts were identified using the following predetermined hashtags related to infertility: #infertility, #infertilitysuccess, #infertilityhelp, #infertilityawareness, and #infertilitysupport. The search and data collection were performed by the primary investigator, who reviewed 20 posts per day, totaling 500 posts throughout the study period. Posts were sorted by relevance to the searched keywords.

### Inclusion and Exclusion Criteria

Posts were included if they were relevant to the topic of infertility, were in the English language, and contained either text, image, or video content. Repeated entries, posts not primarily focused on infertility, and those not in English were excluded from the study. After applying these criteria, a total of 673 posts were included for analysis.

### Data Analysis

The included posts were analyzed for content relating to various aspects of infertility, such as etiology, prevalence, symptoms, diagnosis, screening, prevention, treatment, mortality, rehabilitation, and support groups. The posts were also categorized based on the type of user who shared them: doctors, health and wellness industry professionals, dieticians, patients with the condition, other allied health specialists, and others.

### Quality and Reliability Assessment

The quality and reliability of the posts were assessed using two validated tools: the DISCERN score and the Global Quality Score (GQS). The DISCERN score is a standardized instrument used to judge the quality of written consumer health information, focusing on the clarity of aims, overall quality of health information (11). Each post was independently assessed by two trained evaluators, and the median scores were calculated for inclusion in the statistical analysis.

### Statistical Analysis

Descriptive statistics were used to summarize the characteristics of the posts, including the type of post (image or video), number of likes, views, and comments, and the distribution of posts across different categories of users and content topics. The quality and reliability scores were analyzed to provide a quantitative measure of the information quality. The data were then divided into two groups for comparative analysis: information posted by healthcare professionals (Group A) and all others (Group B). The Z-test was used to compare the percentage of correct information and the difference in quality and reliability scores between the two groups. A p-value of less than 0.05 was considered statistically significant. Data entry and management were carried out using Microsoft Excel 2020, and SPSS Statistics version 16 was used for the statistical analysis.

### Ethical Considerations

To protect the privacy of individuals, all data were anonymized, and no personal or identifying information was recorded or reported. The study was conducted in compliance with ethical guidelines for research involving social media data.

**Table 1:** Number of relevant posts under each hashtag. This table provides a breakdown of the number of posts analyzed and the number of relevant posts for each specified hashtag.

| Hashtag name | Post analyzed | Relevant posts |
| --- | --- | --- |
| #infertility | 200 | 186 |
| #infertilitysuccess | 220 | 214 |
| #infertilityhelp | 90 | 78 |
| #infertilitysupport | 100 | 89 |
| #infertilityawareness | 110 | 106 |
| Total | 720 | 673 |

**Table 2:**
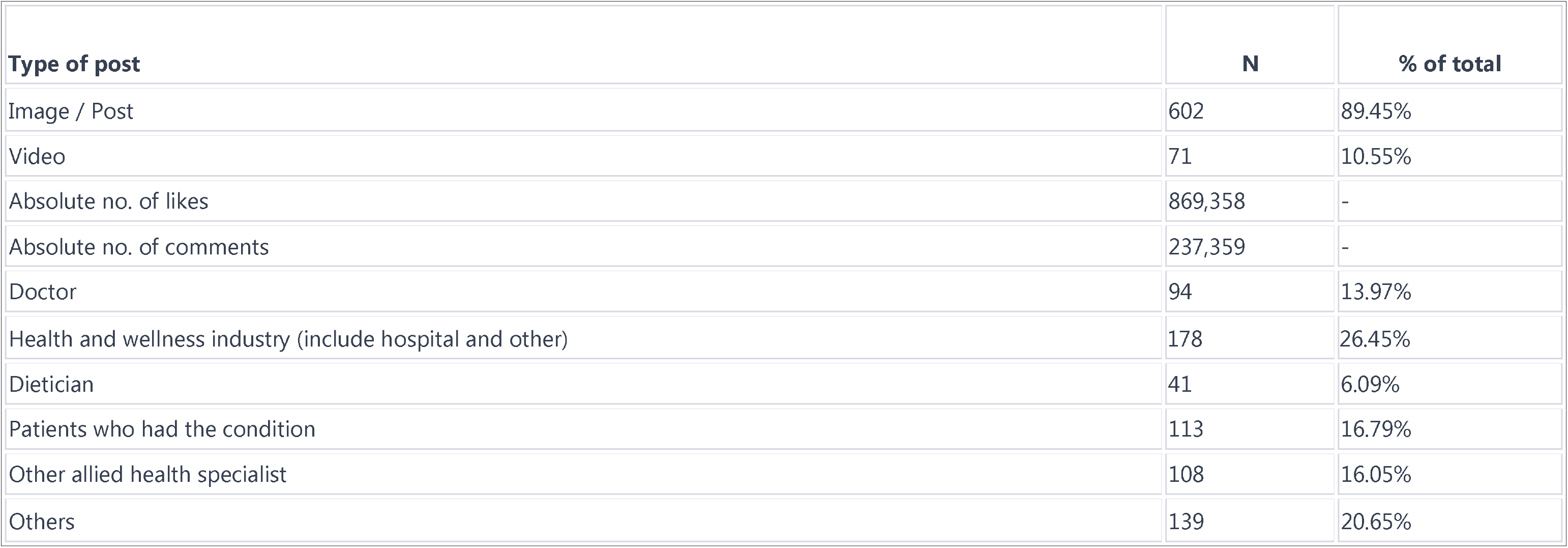
This table provides a detailed breakdown of the type of posts, their count (N), and their percentage of the total, along with the absolute number of likes and comments.

| Type of post | N | % of total |
| --- | --- | --- |
| Image / Post | 602 | 89.45% |
| Video | 71 | 10.55% |
| Absolute no. of likes | 869,358 | - |
| Absolute no. of comments | 237,359 | - |
| Doctor | 94 | 13.97% |
| Health and wellness industry (include hospital and other) | 178 | 26.45% |
| Dietician | 41 | 6.09% |
| Patients who had the condition | 113 | 16.79% |
| Other allied health specialist | 108 | 16.05% |
| Others | 139 | 20.65% |

**Table 3:** This table provides a breakdown of various aspects of the study, including Etiology, Prevalence, Symptoms, and others, along with their respective counts (N) and percentages of the total.

|  | N | % |
| --- | --- | --- |
| Etiology | 316 | 46.95% |
| Prevalence | 403 | 59.88% |
| Symptoms | 425 | 63.15% |
| Diagnosis | 404 | 60.03% |
| Screening | 328 | 48.74% |
| Prevention | 389 | 57.80% |
| Treatment | 506 | 75.19% |
| Mortality | 244 | 36.26% |

**Table 3:**
|  | **N** | **%** |
| --- | --- | --- |
| Rehabilitation | 241 | 35.81% |
| Support groups | 297 | 44.13% |

**Table 4:** Quality and reliability of the posts. This table provides a detailed breakdown of the quality and reliability of the posts, categorized into different levels from very low to very high, along with their respective counts (N) and percentages of the total.

|  | **Subcategory** | **N** | **%** |
| --- | --- | --- | --- |
| Global Quality Score | 1 Very low (add sentence) | 37 | 5.50% |
|  | 2 Low | 76 | 11.29% |
|  | 3 Medium | 272 | 40.42% |
|  | 4 High | 95 | 14.12% |
|  | 5 Very High | 193 | 28.68% |
| Reliability Score | 1 | 29 | 4.31% |
|  | 2 | 89 | 13.22% |
|  | 3 | 254 | 37.74% |
|  | 4 | 102 | 15.16% |
|  | 5 | 199 | 29.57% |

**Table 5:** This table compares Group A and Group B in terms of the number of correct posts and the percentage of correct posts, along with the P value indicating the statistical significance of the difference between the groups.

|  | **Group A (n= 382)** | **Group B(n=291)** |
| --- | --- | --- |
| No of correct post | 361 | 196 |
| Percentage | 94.50 | 67.01 |
| P value | <0.05 | - |

**Table 6:** This table provides a comparison between Group A and Group B in terms of the Global Quality Score and Reliability Score, detailing the mean and standard deviation for each group along with the P value indicating the statistical significance of the differences.

|  | Group A (382) | Group B (291) | P value |
| --- | --- | --- | --- |
| Global Quality Score |  |  |  |
| Mean ± SD | 2.28±0.58 | 2.64±0.67 | <0.005 |
| Reliability Score |  |  |  |
| Mean ± SD | 2.57±0.45 | 2.79±0.87 | <0.001 |

## Discussion

The increasing reliance on social media platforms, particularly Instagram, for health-related information has revolutionized the way individuals seek and share experiences about sensitive topics such as infertility. Our cross-sectional observational study analyzed 673 Instagram posts using the DISCERN and GQS scores to understand the quality and reliability of content related to infertility. The findings underscore the critical role of social media in providing a platform for individuals to share and seek information and support, with a particular focus on Instagram’s utility and influence.

Most posts analyzed were images, which resonate with the visual-centric nature of Instagram. This aligns with Barker’s (2008) assertion that electronic support groups and patient-consumer interactions on social media platforms significantly influence health-related decision-making and emotional well-being (12). Our study found that women derive significant benefits from online experiences in coping with infertility challenges, which is consistent with Sormunen et al. (2017)’s findings on the importance of communication and coping strategies among women affected by infertility (13).

However, while the benefits are notable, our study also highlights the risks associated with misinformation and the quality of content shared on information available. This finding is crucial as it underlines the need for healthcare professionals to guide patients towards reliable sources of information and to be proactive in disseminating accurate and helpful content on these platforms, as suggested by the ACOG (2019) (14).

The study also revealed a statistically significant difference in the accuracy of posts between those shared by healthcare professionals and others, emphasizing the need for expert intervention in guiding public discourse on health-related topics. This is supported by Rice and Katz (2002), who discussed the social consequences of internet use and the importance of access to accurate information (15).

One of the most poignant findings of our study is the emotional support and community building that Instagram facilitates for individuals dealing with infertility. Patients often shared personal journeys and sought support from others undergoing similar experiences. This observation is in line with Jones et al. (2020)’s research, which highlighted the importance of online fertility educational materials and the perceived benefits by fertility patients (16). The emotional resonance and shared experiences provide a unique form of support that is not readily available in traditional healthcare settings.

Furthermore, the study delved into the types of content shared by different users. Patients were more inclined to share personal experiences and emotional narratives, especially related to IVF treatments. In contrast, healthcare professionals focused on educational content and promoting informed decision-making, particularly regarding oocyte cryopreservation. This dichotomy in content types underscores the multifaceted nature of social media platforms in health communication, as discussed by Moorhead et al. (2013) (17).

The rise in patient inquiries about cryopreservation and the prevalence of promotional content from fertility centers and direct-to-consumer fertility testing companies raise questions about the commercialization of health-related information on social media. The competition among fertility centers and the growth of these companies indicates a market-driven aspect of health communication on platforms like Instagram, which users need to be aware of.

In the context of the broader digital healthcare revolution, our findings align with the World Health Organization’s efforts to leverage eHealth programs to remove barriers to health and provide extensive benefits for healthcare. The internet, particularly social media, is increasingly being used as a tool for health communication and information dissemination.

However, study is not without its limitations. The reliance on the DISCERN and GQS scores, while providing a structured approach to assessing content quality, may not capture all nuances of the posts. Additionally, the cross-sectional nature of the study limits the understanding of the long-term impact and changes in content over time.

In conclusion, our study highlights the significant role of Instagram in facilitating discussions and support networks around infertility. The platform provides a unique space for emotional support, information sharing, and community building. However, the quality and reliability of the content vary, necessitating a proactive role from healthcare professionals in guiding patients and the public towards accurate information. Future research should focus on longitudinal studies to understand the evolving nature of health-related content on social media and explore

## Conclusion

Our study delves into the multifaceted role of Instagram in the infertility discourse, examining 673 posts to uncover how individuals, particularly women, seek solace, support, and information. We’ve found that while Instagram serves as a vital platform for emotional support and community building, the reliability and quality of information present significant concerns. The influx of promotional content often muddies the informational waters, highlighting the necessity for healthcare professionals to guide their patients towards credible sources. This study underscores the importance of healthcare providers not merely observing but actively engaging in these digital spaces to combat misinformation and enhance the quality of online health discussions. As the digital age continues to mold the landscape of health communication, Instagram stands out as a pivotal platform in the narrative of infertility, offering a double-edged sword of community support and potential misinformation. The imperative role of healthcare professionals in ensuring a balanced, informed, and empathetic online environment is more crucial than ever. Our findings advocate for a judicious approach to leveraging social media’s benefits while mitigating its risks, aiming to empower those on their journey through infertility with accurate and supportive resources.

## Data Availability

All data produced in the present study are available upon reasonable request to the authors

